# Food & You: A Digital Cohort on Personalized Nutrition

**DOI:** 10.1101/2023.05.24.23290445

**Authors:** Harris Héritier, Chloé Allémann, Oleksandr Balakiriev, Victor Boulanger, Sean F. Carroll, Noé Froidevaux, Germain Hugon, Yannis Jaquet, Djilani Kebaili, Sandra Riccardi, Geneviève Rousseau-Leupin, Rahel M. Salathé, Talia Salzmann, Rohan Singh, Laura Symul, Elif Ugurlu-Baud, Peter de Verteuil, Marcel Salathé

**Affiliations:** Digital Epidemiology Lab, School of Life Sciences, School of Computer and Communication Sciences, EPFL, Switzerland; Department of Statistics, Stanford University, USA

**Keywords:** personalized nutrition, digital cohort, gut microbiota, glycemia

## Abstract

Nutrition is a key contributor to health. Recently, several studies have identified associations between factors such as microbiota composition and health-related responses to dietary intake, raising the potential of personalized nutritional recommendations. To further our understanding of personalized nutrition, detailed individual data must be collected from participants in their day-to-day lives. However, this is challenging in conventional studies that require clinical measurements and site visits. So-called digital or remote cohorts allow *in situ* data collection on a daily basis through mobile applications, online services, and wearable sensors, but they raise questions about study retention and data quality. “Food & You” is a personalized nutrition study implemented as a fully digital cohort in which participants track food intake, physical activity, gut microbiota, glycemia, and other data for two to four weeks. Here, we describe the study protocol, report on study completion rates, and describe the collected data, focusing on assessing their quality and reliability. Overall, the study collected data from over 1000 participants, including high-resolution data of nutritional intake of more than 46 million kcal collected from 315,126 dishes over 23,335 participant days, 1,470,030 blood glucose measurements, 49,110 survey responses, and 1,024 stool samples for gut microbiota analysis. Retention was high, with over 60% of the enrolled participants completing the study. Various data quality assessment efforts suggest the captured high-resolution nutritional data accurately reflect individual diet patterns, paving the way for digital cohorts as a typical study design for personalized nutrition.

## Introduction

Nutrition plays a significant role in moderating the risk and/or severity of several diseases, such as type 2 diabetes^1^, cardiovascular diseases,^2,3^ or cancer^4^. Findings from nutritional epidemiology studies have led to dietary guidelines and public health campaigns designed to support healthy diets. However, while these recommendations are generally based on results aggregated at the population level, a more individualized approach to health^5^ has led to the concept of personalized nutrition. For example, a randomized study showed that personalized recommendations improved diet quality as measured by the Healthy Eating Index (HEI) compared to a control group^6^. Zeevi and colleagues showed how personalized nutrition algorithms could be used to design diets that lower postprandial glucose responses^7^. Further studies showed the importance of personal features such as gut microbiota compositions on glycemic responses^8,9^. Another intervention study showed significant improvement in the food categories consumed when receiving personalized diet advice, compared to generic or no advice^10^. These findings highlight the need for a more holistic and individual approach to nutritional epidemiology, encompassing diet, gut microbiota, physical activity, and lifestyle.

Blood glucose response is a particularly interesting outcome measure for nutritional studies due to its association with insulin resistance, metabolic syndrome, and diseases such as stroke, type 2 diabetes, and heart disease^11^. Reducing blood glucose levels is thus a recommendation of public health authorities around the globe. Identified risk factors for elevated blood glucose levels include a carbohydrate-rich diet, lack of physical activity, and poor sleep, among others. In addition, studies have begun to investigate the role of the gut microbiome in modulating the blood glucose response to food intake. Nutritional studies trying to understand postprandial glucose response (PPGR) are thus faced with the challenge of obtaining data on relevant factors all at once, ideally continuously and *in situ*, that is, in the regular environment in which participants’ lives unfold.

In digital health studies - also called remote or siteless studies - all interactions with participants, as well as data collection, are digital or digitally coordinated. These studies leverage an array of digital devices, wearable sensors, and online services. Digital cohorts and trials have been heralded as a new major development for epidemiological and clinical studies. However, since digital cohorts are a relatively new study approach, open questions regarding selection bias, retention, and data quality remain. Indeed, access to devices connected to the internet and digital literacy may lead to selection bias which in turn may lead to a lack of representativity of the study population compared to the general population. Furthermore, the time burden generated by following the study protocol and collecting the data might create response fatigue, which could in turn translate into lower study adherence, or data quality. Finally, novel data collection methods may not have been thoroughly validated.

Here, we address some of these questions by evaluating the completion rates, adherence, and data quality of the “Food and You” digital cohort on personalized nutrition. The “Food & You” study started in early 2019 and consisted of two distinct digital sub-cohorts; the sub-cohort “Basic” (cohort B) restricted to non-diabetic participants, and the sub-cohort “Cycle” (cohort C) restricted to non-diabetic women of reproductive age who did not use hormonal contraceptive or medication (see Methods for inclusion / exclusion criteria). The study duration for cohort B was 14 days, whereas cohort C participants were enrolled for 28 days, matching the length of a typical menstrual cycle. The study was performed in Swizterland, and all participant were required to have a postal address in Switzerland. Throughout the study, participants were requested to report i) food consumption using an AI-assisted food tracking app (MyFoodRepo), ii) continuous blood sugar levels using a continuous glucose monitor, and iii) physical activity and sleep using either activity trackers or daily surveys. They were furthermore asked to follow a protocol that included a one-time stool sample collection for gut microbiota analysis, and the consumption of standardized breakfasts.

The present paper details the study protocol, and reports study engagement data by looking at the individual characteristics of participants on their journey from enrollment to completion. Further, we provide an overview of the data collected in the “Food & You” cohort from October 2018 to March 2023, and describe our efforts to assess data quality, including the comparison of nutritional and microbiota data collected in “Food & You” with data collected in traditional (on-site) studies. We also discuss the challenges of running a complex digital cohort, and how we addressed them. Overall, retention rates were relatively high, with more than 60% of enrolled participants completing the study. Despite certain fatigue over time, adherence very high, especially for glucose response data, nutritional data, microbiota data, and data from daily surveys. While the study population shows some demographic differences compared to the overall population, the nutrition patterns are in very good agreement with data obtained in another study from a representative sample of the general Swiss population.

## Methods

### STUDY DESIGN AND SETTING

The Swiss “Food & You” study is a digital cohort study collecting data on glycemia, nutrition, gut microbiota, lifestyle, and physical activity as well as demographic data (Figure 1). The cohort was open to anyone fulfilling the inclusion criteria listed in Supplementary Table 1. The study consisted of four sequential phases: enrollment phase, preparatory phase, tracking days phase, and follow-up phase (Figure 2). In the *enrollment phase*, interested participants were first required to perform a self-check of their eligibility, and fill out a consent form and a short screening questionnaire. Following this, and upon acceptance by a member of the study team (based on the available capacity to accommodate new participants), the participants were considered enrolled. During the subsequent *preparatory phase*, enrolled participants were given instructions on the “Food & You” website and asked to fill out a series of questionnaires. Next, they were instructed to download the AI-assisted nutrition tracking app MyFoodRepo^1^ (MFR) to track their food intake for a trial period of at least three days. After the successful completion of the trial period, participants would order the study material which included, among other things, a continuous glucose monitoring (CGM) sensor for each period of 14 days and stool-sample self-collection material. Upon receipt, they were asked to choose the starting date of their tracking days phase. In the *tracking days phase*, participants were required to log all their food and drink intake via the MyFoodRepo app, wear the CGM sensor, and answer two daily surveys for 14 (cohort B) and 28 (cohort C) days. In addition, participants were instructed to take standardized breakfasts in accordance with dietary restrictions from the 2nd to the 7th day during the first week. They were asked to avoid altering standardized meals, as well as to refrain from eating or engaging in physical activity for the subsequent two hours. Cohort C participants repeated these instructions on their third tracking week (Supplementary Table 2). On days 6 and 7 (additionally on days 21 and 22 for cohort C) they were asked to perform an oral glucose tolerance test by drinking 50g of glucose. In addition, study participants were asked to provide one stool sample collected anytime during the tracking days phase. At the end of the tracking days phase, participants were asked to upload their physical activity and CGM data on the “Food & You” website. In the *follow-up phase*, they were requested to fill out a feedback questionnaire regarding their experience. Participants were also provided with interactive visualizations of their data (Supplementary Figure 1). Cohort C participants were followed for two additional menstrual cycles during which they continued to track their menstrual cycle and fertility-related body signs such as morning temperature and cervical mucus characteristics.

**Figure 1.**
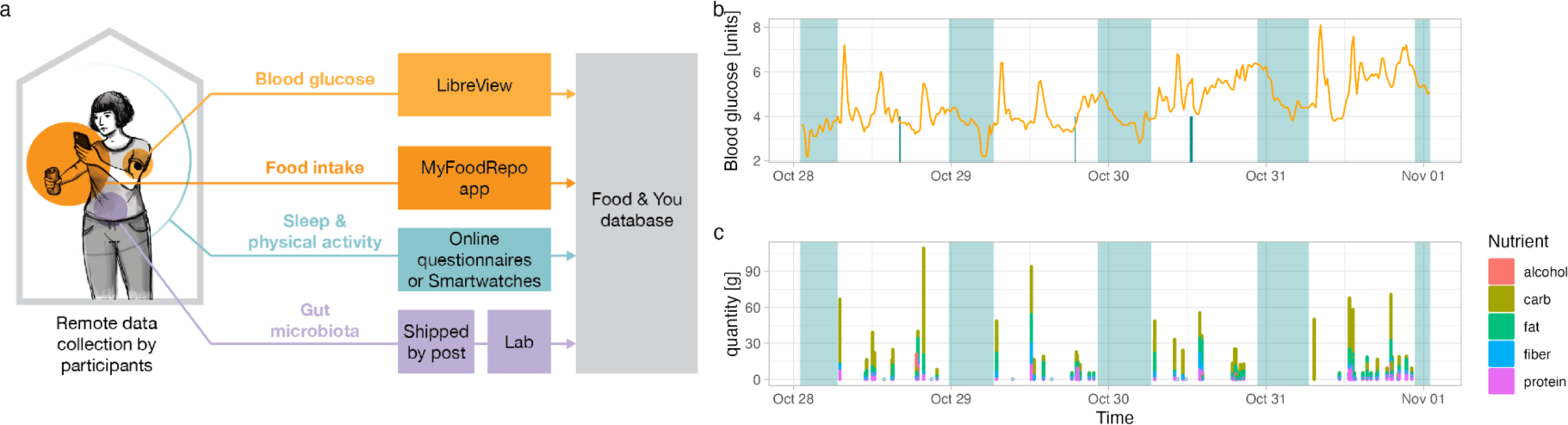
Data collection. a) Schematic illustrating the data collection process. (Left) Participants track the study variables *in situ* (from home, work, etc.). (Center) Data or samples are collected via web platforms and apps, or shipped to the lab by mail. (Right) Data is processed in the Food & You database. b and c) Example of data collected by one participant over 5 days. Top panel shows blood glucose levels (orange line), physical activity (turquoise spikes), and sleep (translucent turquoise rectangles). Bottom panel shows time and micronutrient composition (colors) of reported food intake. Like in the top panels, translucent turquoise rectangles show when the participant is asleep.

**Figure 2.**
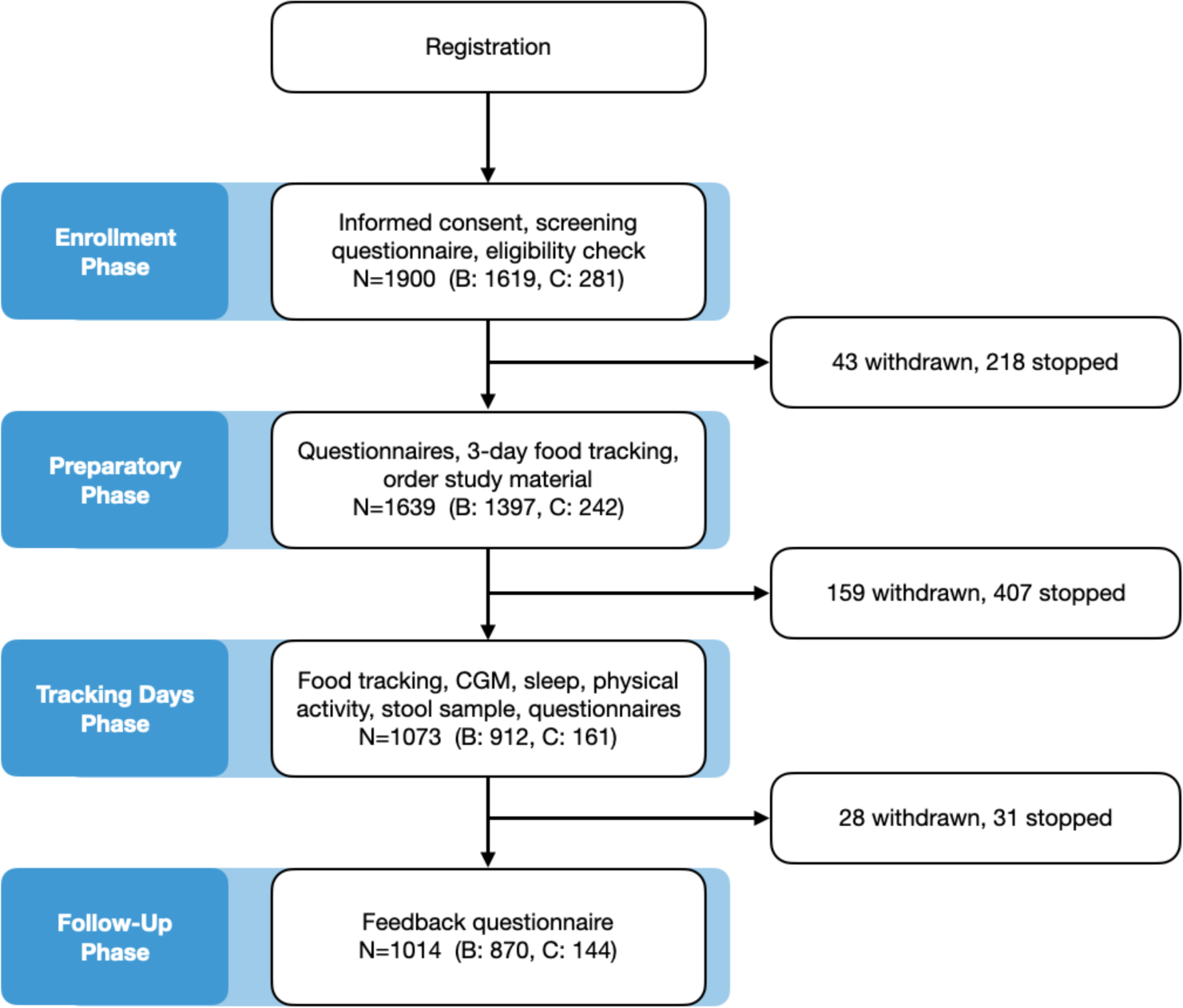
Study phases with participants per phase, and exit numbers.

The Geneva ethics commission has reviewed and authorized the project (Ethical Approval Number: 2017-02124). The study is registered on the website of the Federal Office of Public Health (SNCTP000002833) and the platform clinicaltrials.gov (NCT03848299).

### DATA COLLECTION

#### Questionnaires

During the enrollment phase, interested subjects were requested to fill out a screening questionnaire with items regarding age, gender, height, weight, type of mobile phone and dietary restrictions. During the preparatory phase, enrolled participants had to fill out a lifestyle and health-related questionnaire. Participants were asked about their general health (smoking, diet, food supplement intake, general hunger levels, health state, past diagnoses, antibiotic intake, and menstrual health), physical activity (exercise frequency, duration, and intensity), sleep (bedtime, wake-up time), sociodemographic variables (nationality, socioeconomic status, job status, and household description), and requested to provide self-measured anthropometric measurements (waist and hip circumference, height, and weight). In the tracking days phase, participants had to fill out a short form each evening to validate their adherence to protocol and document medication intake. Cohort C participants had to answer additional questions on menstrual blood, cervical mucus and provide self-reported temperature measurements on a daily basis.

#### Dietary Intake

Participants of the “Food & You” study were asked to log any dietary intake in real time on the MyFoodRepo app (MFR) using one the following options: taking pictures of the food/drink, scanning the product’s barcode (if available), or describing the food item with text. A logged entry is defined as a “dish”, and can contain multiple food items. For example, tuna, steamed potatoes, and green beans are all single food items, and together compose a dish. The pictures were automatically segmented and classified by an image recognition algorithm^12^. Portions, segmentations, and food classes were subsequently verified or edited by a team of trained annotators. The MyFoodRepo app also allows annotators to communicate with the participant for clarifications about the food, and participants were able to leave comments through the app. This process ensured that every single dish in the nutritional data was reviewed by a member of the study team. Each food item was linked to a nutritional value database containing 2’129 items built on the Swiss Food Composition Database^13^, MenuCH data^14^, and Ciqual^15^. When food intake was logged through barcode scanning, nutritional values of the food items were fetched from the Open FoodRepo database API^16^. Manual entries were matched to food items by the annotators.

As the aforementioned nutritional values data sources did not provide standard portion sizes, these were manually extracted from the portion list of the WHO MONICA study^17^, and the Mean Single Unit Weights of Fruit and Vegetables report^18^ by the German Federal Office of Consumer Protection and Food Safety. When a standard portion was not available for a particular food item, we assigned the standard portion of a similar food item.

Each dish logged through the MyFoodRepo app carries a timestamp, which enables dietary analysis at a high temporal resolution. Food items were classified into categories based on the menuCH study^14^ (Chatelan, Marques-Vidal, et al. 2017). Barcoded food items from the Open FoodRepo database were categorized based on the food product description. When such a description was not available, we assigned the category extracted from the Open Food Facts database^2^. While “Food & You” is the first large cohort to use MyFoodRepo, the app annotation quality had previously been validated^19^.

#### Glycemia

Glycemic data was collected by using the Flash Glucose Monitor Freestyle Libre (Abbott Diabetes Care). The system, which has been validated in numerous studies,^20–22^ consists of a disposable sensor applied to the back of a participants’ upper arm, and a reader device or a smartphone app allowing to collect data from the sensor via NFC technology. It measures glycemia every 15 minutes via a subcutaneous filament carrying enzyme glucose sensors^23^. To encourage high adherence to protocol, we chose a non-blinded glucose monitoring system to allow the participants to see their glycemia in real time. Participants self-applied the sensor at home following explanations provided in writing and video. Cohort B participants wore a single sensor for 14 days, whereas cohort C participants wore two sensors consecutively for a period of 28 days to cover the length of a typical menstrual cycle. Notably, when participants scan the sensor, the data from the previous eight hours is collected. Thus, unless participants scan the sensor at least every 8 hours, some data may remain unretrievable.

#### Gut Microbiota

Participants were requested to collect a stool sample following detailed written and video instructions. They could collect and ship their sample anytime during the tracking days phase. Samples were collected with stool nucleic acid collection and preservation tubes from Norgen Biotek, stored at room temperature and shipped in batches of 100 to 192 samples to Microsynth AG (Balgach, Switzerland) for sequencing and bioinformatics analysis. V4 region of the bacterial 16S rRNA gene was sequenced via creation of two-step Nextera PCR libraries using the primer pair 515F (NNNNNGTGYCAGCMGCCGCGGTAA) and 806R (NNNNNGGACTACNVGGGTWTCTAAT). The primers use 5 bases at their 5’ end to increase diversity of the bases during the first five sequencing cycles. Subsequently, the Illumina MiSeq platform and a v2 500 cycles kit were used to sequence the PCR libraries. The produced paired-end reads which passed Illumina’s chastity filter were subject to de-multiplexing and trimming of Illumina adaptor residuals using Illumina’s real time analysis software included in the MiSeq reporter software v2.6 (no further refinement or selection). The quality of the reads was checked with the software FastQC version 0.11.8. The locus specific V4 primers were trimmed from the sequencing reads with the software cutadapt v2.8. Paired-end reads were discarded if the primer could not be trimmed. Trimmed forward and reverse reads of each paired-end read were merged to in-silico reform the sequenced molecule considering a minimum overlap of 15 bases using the software USEARCH version 11.0.667. Merged sequences were then quality filtered allowing a maximum of one expected error per merged read. Reads that contained ambiguous bases or were considered outliers regarding the amplicon size distribution were also discarded. Samples that resulted in less than 5000 merged reads were discarded, to not distort the statistical analysis. The remaining reads were denoised using the UNOISE algorithm^24^ implemented in USEARCH to form amplicon sequence variants (ASVs) discarding singletons and chimeras in the process. The resulting ASV abundance table was then filtered for possible bleed-in contaminations using the UNCROSS^25^ algorithm, and abundances were adjusted for 16S copy numbers using the UNBIAS^26^ algorithm. ASVs were compared against the reference sequences of the RDP 16S database, and taxonomies were predicted considering a minimum confidence threshold of 0.5 using the SINTAX algorithm^27^ implemented in USEARCH.

#### Physical Activity and Sleep

Study participant’s physical activity (PA) and sleep data were collected using one of two methods: objectively via Apple Health, Google Fit, or smart-watches, or subjectively, i.e. self-reported on the study website via the morning and/or evening questionnaire. The different formats of the objective PA and sleep data were harmonized and stored in a single database, and comprised daily step count, daily calories burned, bedtime, and wake-up time. In addition, for PA measured with smart devices, the type of physical activity, the start and end times, the amount of burned calories, the average heart rate, and the maximum heart rate were collected. Participants self-reporting their sleep and PA on the website had to report the times at which they fell asleep and woke up, if they did any physical activity, and if so, when they started and finished, as well as the perceived intensity of their effort.

### STATISTICAL ANALYSIS

#### Data Preproccessing

Sociodemographic questionnaires variables and anthropometric measurements were re-coded as follows; Monthly household income was classified in 4 categories: <6000, 6000 to 8999, 9000 to 12999 and >=13000 Swiss Francs (CHF). Note that the median income in Switzerland was 6,665 CHF per month in 2020. Household type categories were defined as either “alone” for single-person households, “couple w. children”, “couple w.out children”, and “other”. Age at study start was transformed into the following categories; 18 to 34, 35 to 49, 50 to 64, and older than 65 years. Citizenship was categorized as “Swiss”, “Binational”, or “Foreigner”. Education level was categorized as “low” (mandatory, primary school), “intermediate” (high school and professional diploma) or “high” (university). Smoking status was categorized as “non smoker” (never smoked), “former smoker” or “current smoker” (occasionally or daily). The self-rated health questions were re-coded as binary variables “Not good to average” (not good, average, somewhat good) or “Good to very good” (good, very good). Residential addresses were geocoded, and municipality number (Spatial Development ARE) was extracted for each address. The municipality number was then joined with a urban/rural municipality typology and linguistic region dataset from the Swiss Federal Statistical Office so that each address was categorized as “urban” or “rural,” whereas linguistic region was categorized as “german” or “latin,” the latter encompassing french, italian, and romansh-speaking regions.

Missing values in income and education (<0.2% and less than 8%, respectively) were imputed via multiple imputations with chained equations using the mice package^28^. Self-reported general physical activity levels were assessed based on the weekly frequency and average duration in minutes. The data was then coded into “active” and “inactive” based on the WHO cutoff of 75 min per week^29^. Body-mass index (BMI) was calculated based on self-reported height and weight and classified into “Underweight,” “Normal,” “Overweight,” “Obese” based on the WHO cutoffs for BMI. For food intake, we removed days for which energy intake was below 1’000 kcal and aggregated by study subject to calculate the individual weighted mean to account for day-of-week and seasonal variations of intake over the observation period.

To assess the extent of glycemic excursions for each participant, we calculated the proportion of readings below, in, and above the target range based on the cut-off values of 3.9 and 10 mmol/L^30^. We also computed the mean amplitude of glycemic excursions (MAGE) for each subject using the R package gluvarpro (4.0).

#### Completion rate and study population analysis

Study completion rate was defined as the ratio of the number of participants having completed the study over the total number of participants enrolled in the study. The completion rate was then calculated for each cohort (B and C) and each of the following strata: gender, age group, BMI, phone type, and dietary restriction.

For the study population analysis, we only included the participants who have completed the study, and calculated their proportion in each cohort for the following strata: gender, household income, household type, age group, citizenship, education, smoking status, health status, urbanity, linguistic region, physical activity, and BMI.

Analysis of adherence to study protocol was conducted by cohort given that their tracking phase duration differed. For each participant, we reported the number of days with (i) CGM measurements, (ii) reported food (distinguishing between days with a total intake above 1000 kcal or days with any food logged), (iii) sleep data, (iv) PA data, and (v) questionnaire data. We also reported the number of standardized breakfasts taken by participants, of glucose oral tolerance tests performed, and stool samples collected.

#### Data quality analysis

Given the multimodal nature and multi-dimensionality of the collected data, assessment of data quality may take several forms. Here we performed a series of analyses to evaluate if each type of collected data followed expected patterns.

Specifically, we performed three analyses assessing the food data quality with respect to timing, adherence, and composition. First, we assessed the distributions of food intake times by calculating the count of the logged food intakes by day of the week and hour of the day. Second, we hypothesized that most CGM peaks should be preceded by a food intake. We thus conducted an experiment using CGM data as ground truth. For each CGM peak (*i.e*., a local maximum above the 90th percentile of the participant’s CGM distribution) we reported whether a food intake of at least 50 kcal had been logged within the 90 min interval preceding the peak, or within the 90 min preceding the peak minus 2 hours (control case). We then reported, for each participant, the proportions of CGM matching a food intake in both experiment and control situations. To test our hypothesis that the proportion of peaks with logged food intake within 90 minutes of the peak would be higher than the same proportion for the control time-window, we used a non-parametric Wilcoxon rank sum test. Third, to assess the quality of the composition of reported food intake, we compared food intake data obtained in the “Food & You” cohort to data reported by the Swiss nutritional survey menuCH which was obtained from a demographically representative sample. We considered amounts of energy, meat, dairy, water intake as well as the proportion of the study population that consumed more than five portions of fruits and vegetables a day. For each of the following strata: sex, age group and linguistic region, we calculated the weighted means of the intake to account for weekday variations.

For sleep, the data were aggregated at the participant level to calculate the weekday and weekend individual mean values for bedtime, wake-up time, and sleep duration. Sleep durations of less than 4 hours or more than 12 hours were excluded.

For gut microbiota data comparison, we used relative abundances for the microbiome samples of other studies available from the R package CuratedMetagenomicsData (version 3.7). These samples were selected based on the filtration criteria that all samples were of gut origin and from healthy adults. Four studies with most of such samples were selected for comparison with the “Food & You” microbiome^7,31,32 33^. Relative abundances from these studies and “Food & You” were aggregated at the genus taxonomic level. Bray-Curtis distances were calculated between the samples using the vegdist function of the vegan package in R, which were then transformed into two-dimensional principal coordinates using the pcoa tool of ape package in R.

## Results

### STUDY COMPLETION RATE

Overall study completion rate - defined as the ratio of the number of participants having completed the study over the total number of participants enrolled in the study - was relatively stable over the years, with 69.5%, 56.4%, 65.7%, and 57.3% in 2019, 2020, 2021, and 2022, respectively. Detailed completion rates are shown in Table 1. Within the investigated groups - gender, BMI, age, phone type, and diet - we did not see any major differences, with the exception of diet. Participants who reported dietary restrictions had higher completion rates (82.5% and 81.5% in cohorts B and C, respectively) than their counterparts without restrictions (62.7% in cohort B and 56.7% in cohort C).

**Table 1.** Cohort completion rates. For each section, the table shows percentages per section and cohort, and absolute numbers in parentheses. Percentages are calculated as the number of participants having completed the study over the number of participants enrolled in the study.

|  |  | **Cohort B** | **Cohort C** |
| --- | --- | --- | --- |
| Age Group | 18-34 | 63.27 (348) | 61.76 (84) |
|  | 35-49 | 64.23 (316) | 59.18 (58) |
|  | 50-65 | 60.4 (183) | 50.0 (2) |
|  | 65+ | 57.5 (23) | 0.0 (0) |
| BMI | Normal | 63.64 (574) | 62.15 (110) |
|  | Underweight | 61.54 (16) | 63.64 (7) |
|  | Overweight | 63.4 (220) | 47.62 (14) |
|  | Obese | 57.14 (60) | 58.33 (7) |
| Gender | female | 63.24 (425) | 59.5 (144) |
|  | male | 61.38 (445) | 0.0 (0) |
| Dietary Restriction | No | 62.73 (771) | 56.74 (122) |
|  | Yes | 82.5 (99) | 81.48 (22) |
| Phone Type | Android | 66.44 (396) | 56.14 (64) |
|  | iPhone | 59.18 (474) | 62.5 (80) |

### COHORT CHARACTERISTICS

1,014 study subjects have completed the “Food & You” study, of which 870 in cohort B and 144 in cohort C (Table 2). In cohort B, both sexes were equally distributed. The proportion of healthy and educated subjects was found to be higher in “Food & You” as compared to the Swiss population. A direct comparison is difficult, because representative statistics for the Swiss subpopulation with study inclusion and exclusion criteria applied are not available. For example, in cohort B, the proportions of overweight and obese cohort participants was 24% and 6%, while these proportions are slightly higher in the general Swiss population (31% and 11% respectively). However, the combination of diabetes as an exclusion criterion and the well-known association between diabetes and overweight / obesity^34^ may explain the difference. Because representative data from the Swiss population filtered by our exclusion and inclusion criteria is not available, we did not test whether the observed differences could be due to chance.. Some of the large differences are unlikely to be explained by exclusion and inclusion criteria alone. Most strikingly, the “Food & You” study population is much more highly educated, with almost three quarters of participants having a university degree (compared to less than one third in Switzerland). In addition, the 18-34 and 45- to 49 age classes are substantially over-represented, while the elderly (ages 65 and over) are severly underrepresented (less than 3% of participants).

**Table 2.** Cohort characteristics. For each section, the table shows percentages per section and cohort, and absolute numbers in parentheses. Household income is monthly in Swiss Francs (CHF), which is roughly in parity with USD.

|  |  | **Cohort B (n)** | **Cohort C (n)** |
| --- | --- | --- | --- |
| Gender | Female | 48.85 (425) | 100 (144) |
|  | Male | 51.15 (445) | 0 (0) |
| Household Income | <6000 | 17.82 (155) | 22.92 (33) |
|  | 6000-8999 | 20.11 (175) | 24.31 (35) |
|  | 9000-12999 | 29.31 (255) | 28.47 (41) |
|  | >=13000 | 32.76 (285) | 24.31 (35) |
| Household Type | Alone | 18.28 (159) | 21.53 (31) |
|  | Couple w. Children | 30.69 (267) | 33.33 (48) |
|  | Couple w.out Children | 35.06 (305) | 29.17 (42) |
|  | Other | 15.98 (139) | 15.97 (23) |
| Age Group | 18-34 | 37.82 (329) | 57.64 (83) |
|  | 35-49 | 35.86 (312) | 40.97 (59) |
|  | 50-64 | 23.56 (205) | 1.39 (2) |
|  | 65+ | 2.76 (24) | 0 (0) |
| Citizenship | Swiss | 52.64 (458) | 45.14 (65) |
|  | Binational | 10.69 (93) | 13.19 (19) |
|  | Foreigner | 36.67 (319) | 41.67 (60) |
| Education | Low | 1.72 (15) | 2.78 (4) |
|  | Intermediate | 24.71 (215) | 15.28 (22) |
|  | High | 73.56 (640) | 81.94 (118) |
| Smoking | Non Smoker | 48.05 (418) | 51.39 (74) |
|  | Former Smoker | 42.18 (367) | 34.03 (49) |
|  | Smoker | 9.77 (85) | 14.58 (21) |
| Health Status | Not Good to Average | 8.85 (77) | 6.25 (9) |
|  | Good to very Good | 91.15 (793) | 93.75 (135) |
| Urbanity | Urban | 76.55 (666) | 78.47 (113) |
|  | Rural | 23.45 (204) | 21.53 (31) |
| Linguistic Region | German | 47.93 (417) | 53.47 (77) |
|  | Latin | 52.07 (453) | 46.53 (67) |
| Physical Activity | Active | 66.55 (579) | 65.97 (95) |
|  | Not active | 33.45 (291) | 34.03 (49) |
| BMI | Underweighted | 1.84 (16) | 4.86 (7) |
|  | Normal | 65.98 (574) | 76.39 (110) |
|  | Overweighted | 25.29 (220) | 13.89 (20) |
|  | Obese | 6.9 (60) | 4.86 (7) |

### ADHERENCE

Of the participants who completed the study, adherence to protocol was generally high, with a large majority of participants able to collect the requested amount of data for most modalities. In cohort B, 93.9% provided at least 13 days of food tracking data with daily energy intake above 1000 kcal, whereas this figure was slightly lower (84.7%) for cohort C participants, albeit for at least 27 days. Cohort C participants were asked to provide self-reported mucus quality and temperature measurements on a daily basis. 97% and 76% reported bleeding data on any day, and during the last week, respectively. The 1,014 participants who completed the study logged a total of 297,626 dishes amounting to 43.6 million kcalk. Participants mostly logged their food intake by taking pictures (76.1%), whereas a smaller proportion of entries were logged by barcode scanning (13.3%) or manually (10.6%). Sleep data was available for 64.8% of the participants who completed the study. In total, participants ate 6,944 standardized breakfasts including 2,158 glucose drinks.

In total, 6’460 subjective (i.e. self-reported) physical activities were reported. A large proportion (33% in cohort B, 42.4% in cohort C) of users in both cohorts did not report any activity at all, but the proportion that did report activities was higher in cohort C (Figure 3). Around 4/5 of the participants who provided physical activity data reported only subjective activity via website questionnaire. In terms of objective activities collected through activity trackers such as smartwatches, the most reported activity types (66.5%) were walking, running, and cycling.

**Figure 3.**
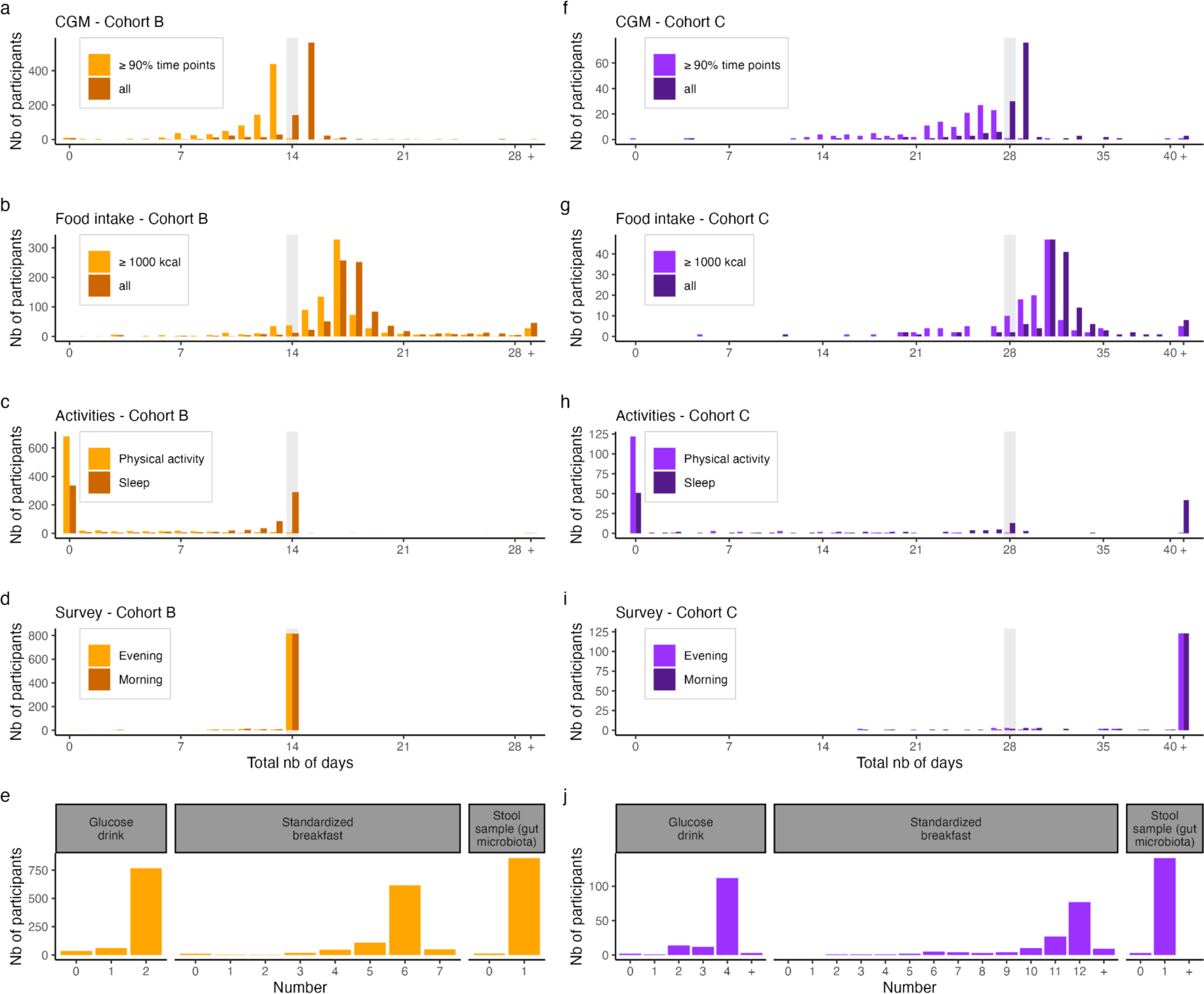
Distribution of collected data. (a-d) and (f-i): Distribution of the total number of days (x-axis) with collected data by participants (y-axis) in cohort B (a-d) and cohort C (f-i) for each of the study data modality. Light gray vertical bar indicates the duration of the study for both cohorts (i.e., the number of days for which participants were instructed to report data). Top panels (a,f) show that distribution for the blood glucose data (CGM: continuous glucose monitoring). Darker shades show the distribution in the case where days are included if there is at least 1 data point collected that day, while lighter shades show the distribution in the case that days are included only if there are data points for at least 90% of the day. 2nd row panels (b, g) show the distribution for the food intake data. In a similar fashion to the top panels, darker shades show the distribution for days with any food intake, while lighter shades only include days during which total caloric intake was above 1000 kcal. 3rd row panels (c, h) show the distribution for the sleep and physical activity data. 4th row panels (d, i) show the distribution for daily questionnaires. Days were included if participants filled the morning (lighter shade) or evening (darker shade) surveys. 5th row panels (e, j) show the number of glucose drink (left), standardized breakfast (center), and stool samples (right) reported or sent by participants. Cohort B (resp. C) participants were instructed to eat two (four) glucose drinks and 6 (12) standardized breakfasts. For panel (i) and (h), note that participants in Cohort C were allowed to keep answering survey for 150 days after start.

In total, 997 participants who completed the study provided a stool sample for microbiota composition quantification. The distribution of relative abundances at phylum and species levels are depicted in supplementary figures 2 and 3.

### DATA QUALITY

Breakfasts were observed from 05:00 onwards, whereas the hourly distribution of this meal was shifted later during the weekend as compared to weekdays (Figure 4a). Logged lunches were found to be consistent with work schedules during the week, with a peak at noon, whereas the number of entries at those hours was much lower during weekends. The number of entries after 20:00 tended to be higher Fridays and Saturdays, likely reflecting social dinners. In general, and as expected, a shift in the later hours was observed for all meals taken during the weekend as compared to weekdays (difference in average meal times of 33 minutes, Wilcoxon rank sum test p-value < 0.001). The exception to this pattern was Sunday dinners: the timing of those was more similar to Monday dinners’ than to Saturday dinners’.

**Figure 4.**
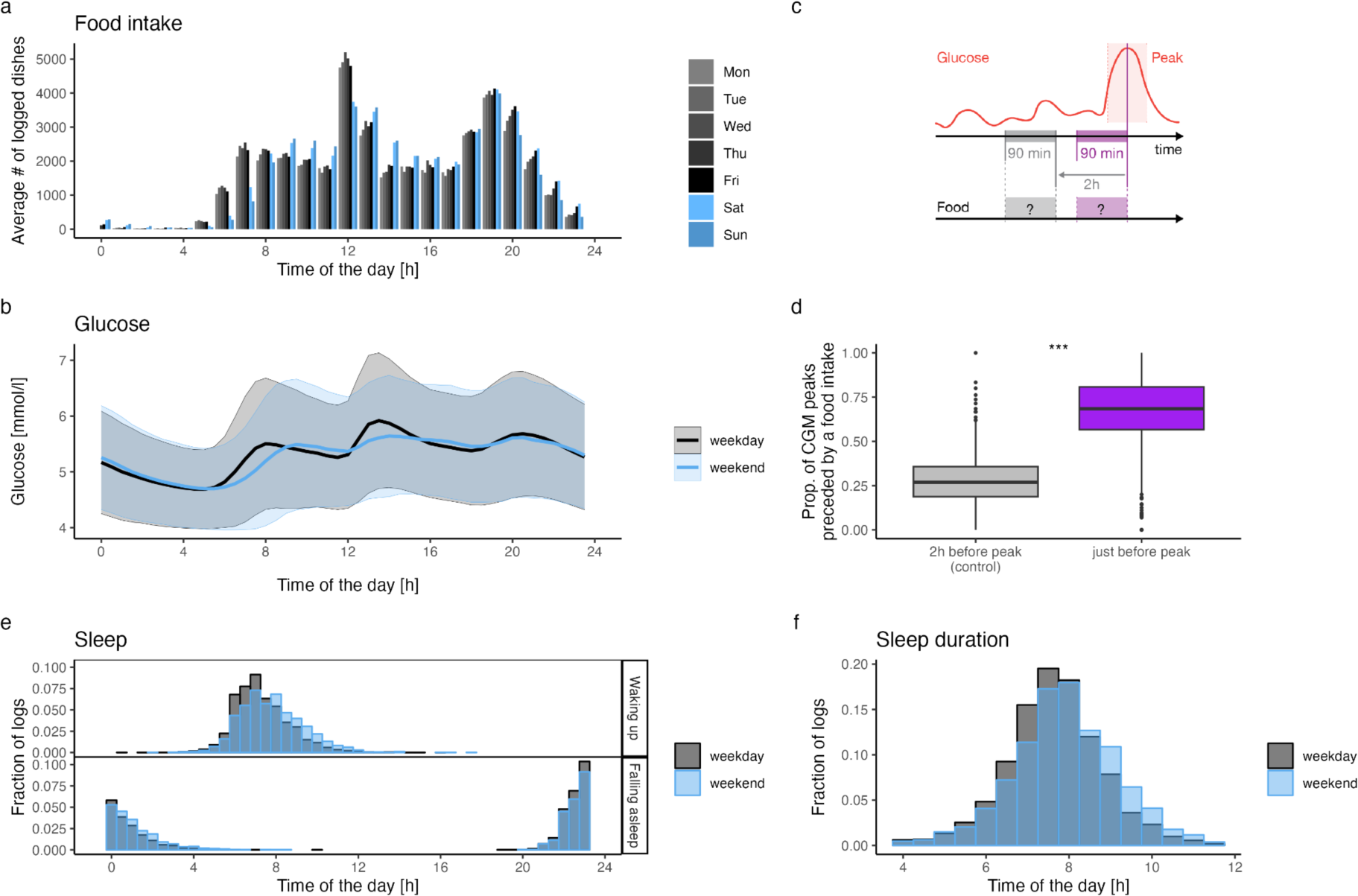
Temporal patterns of measured or self-reported data. a. Total number across all participants of logged dishes (any food or drink intake) per hour (x-axis) and weekday. b. Mean (solid line) and 50% interquartile range (shaded areas) of the blood glucose levels measured during weekdays (black) or weekends (blue). c. Schematic illustrating the experiment to assess if glucose peaks are more likely to be directly preceded by food intake. The time-window of interest (*i.e.*, in which food intake is expected) is displayed in purple, while the control time-window is displayed in gray. d. Distribution of the percentage of glucose peaks directly (purple) or distantly (control - gray) preceded by food intake. Percentages are computed per participant: for each participant, glucose peaks are identified, food intake in the two time-window is reported as a binary variable, and percentages are computed as the fraction of time-windows with reported food intake. e. Distribution of the times at which participants woke up (upper panel) or fell asleep (lower panel) during weekdays (gray) or weekend (blue). These times are either self-reported by participants when filling the morning questionnaires or obtained from participants’ connected devices (smartwatches) data. f. Distribution of average sleep durations (x-axis, in hours) during weekdays (gray) or weekends (blue).

The glucose data displayed a similar pattern as the food data with a clear shift between weekdays and weekend, reflecting the later wake-up and eating times (Figure 4b, maximal cross-correlation between weekend and weekdays mean glucose levels is found with a 30-minute shift). Consistently, wake up and bedtimes were also shifted toward later hours during weekends. Participants slept 7.9 hours a night on average (SD: 1.19, Figure 4f). On average, participants slept 16 minutes longer on weekends than on weekdays (n = 591, paired t-test p-value < 0.001). On average, participants’ bedtime was 23:41 (SD: 1.37,Figure 4e), 23:37 on weekdays, and 23:51 on weekends. Participants woke up on average at 07:35 (SD: 1.43, Figure 4e), and woke up significantly later on weekends, at 07:57 vs 07:27 (average difference of 30 minutes, paired t-test p-value < 0.001).

The proportion of glucose peaks matching a food item was significantly higher (median proportion: 68%) within the experimental group compared to the control group (median proportion: 27%, one-sided Wilcoxon rank sum test p-value < 0.001), indicating a good agreement between the food intake and CGM data. However, as we have no objective ground truth data on nutrition, we do not know what the expected proportions would be.

In general, participants had a healthy glycemic profile with mean amplitude of glucose excursion (MAGE) values of 1.41 mmol/L and 1.21 mmol/L for cohorts B and C, while the proportion of readings above 10 mmol/L (hyperglycemia) was below 0.3%.

### COMPARISON WITH OTHER STUDIES

For participants who completed the study, daily mean energy, meat, dairy and water intake were 2,205.1 kcal, 92 g, 124.9 g, and 964.9 ml, respectively. The proportion of participants more than 5 portions of fruits and vegetables per day was 6.71%. These values were in good agreement with the mean values reported by the Swiss nutritional survey “menuCH” were obtained from a demographically representative sample. The only exception was for water, for which intake was much lower in menuCH than in our study. Means and standard deviations for energy, meat, dairy, water intake as well as proportion of participants eating more than 5 portions of fruits and vegetables per day are reported in supplementary table 3 for the whole population and for various strata (sex, language, age group, and combination of sex and age group).

The PCoA plot of the gut microbiota profile (Figure 5) shows the Bray-Curtis distance dissimilarities of gut microbial community samples found in healthy individuals of “Food & You” and four other gut-related studies. To allow for comparability, bacterial taxonomies from these microbiomes were aggregated at the genus level. The plot shows that most participants from the “Food & You” cohort and the LifeLinesDeep study tend to form their own cluster distinct from other studies.

**Figure 5.**
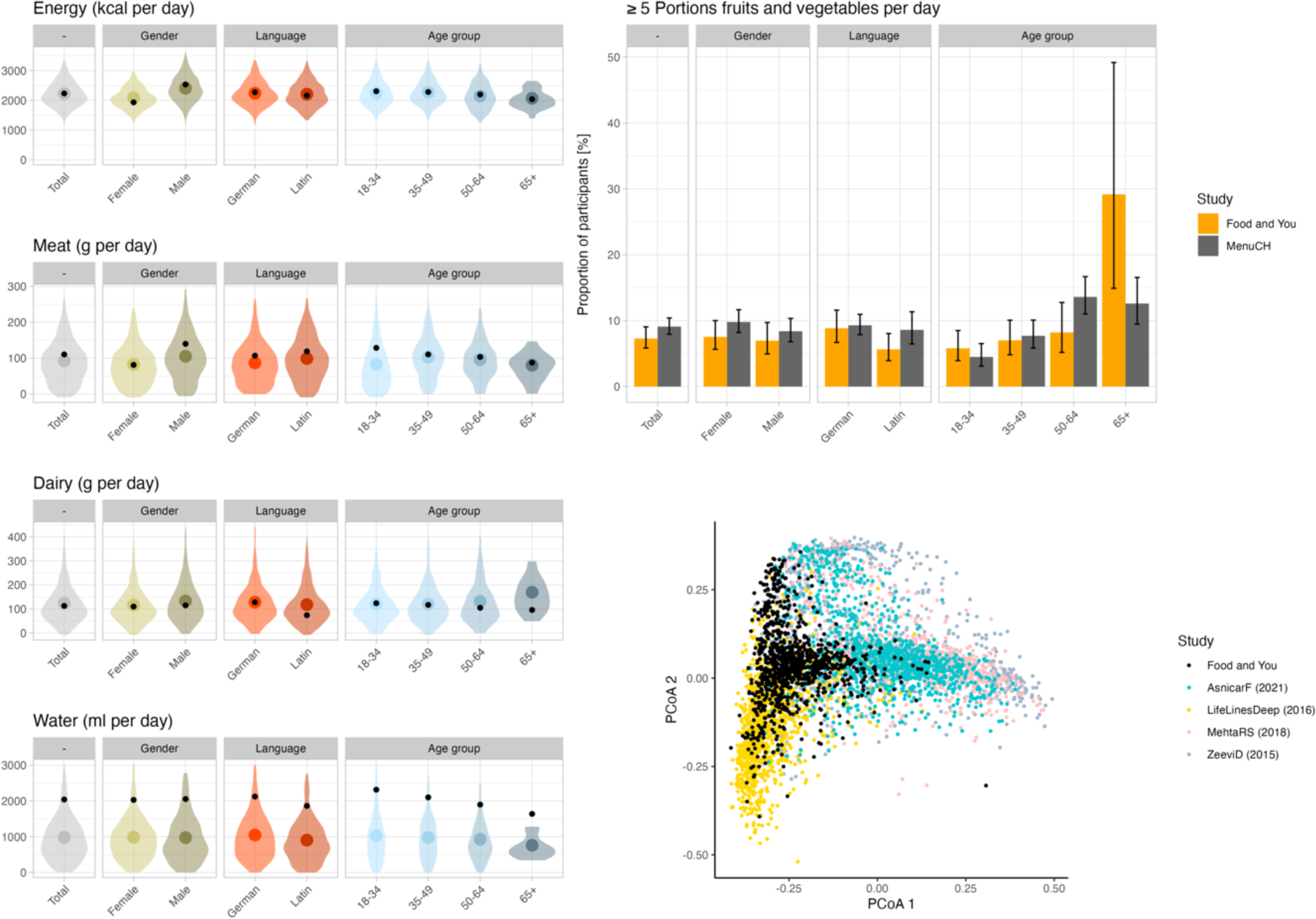
Comparison with other studies. (Left panels): Comparison with the menuCH results. Distribution of the daily intake in energy (top row), meat (2nd row), dairy (third row), and water (bottom row). In each panel, the colored violins and dots represent the distribution and mean of the Food & You data while the smaller black dot is the mean for the corresponding population stratum reported by the menuCH study. (Top right panel): Proportion of participants that, on average, eat more than 5 portions of fruits and vegetables daily. For menuCH, the averages are over 2 days of data collection. For “Food & You”, the average is over 14 (cohort B) or 28 (cohort C) days of data collection. Bar heights show the proportion of participants, black whiskers show the 95% CI. (Bottom right panel): Microbiota composition of F&Y participants compared to that of other cohorts. Each dot is a sample, and samples are colored by cohort. The coordinates of each sample are the first two coordinates resulting from a principal coordinate analysis on the Bray-Curtis distances between samples (Methods).

## Discussion

The “Food & You” study is a fully digital nutrition cohort collecting diverse multimodal data remotely, without any physical contact between the participants and any member of the study team. This study has gathered a wealth of nutritional intake data, blood glucose measurements, survey responses, and gut microbiota samples from 1,014 participants. Here, we described the study protocol, reported on study completion rates, and described the collected data, focusing on assessing their quality and reliability.

The overall completion rate was high, with over 60% of enrolled participants completing the study. In comparison with other digital health studies^35^, the retention rate for 14 and 28 days (cohort B and C, respectively) was rather high. Several factors may have contributed to this outcome. Perhaps most importantly, we approached the study’s design from a participant’s perspective, which led to the decision to develop a new food-tracking app (MyFoodRepo) from scratch, emphasizing ease of use. We also developed the study website and data collection system from scratch in order to directly integrate data collected from sensors or apps on the study website. For example, we combined nutritional data from the app and glucose data from the CGM system to generate interactive charts on the study website. Completion rates in younger and older age groups were comparable, indicating no major hurdle related to the use of digital tools for data collection for older subjects. Subjects with dietary restrictions were particularly committed to the study in both cohorts (completion rates over 80%), in line with previous findings from disease-based digital health studies showing that participants affected by the disease showed higher study retention^35^.

With respect to adherence, data availability was high for most indicators in both cohorts, with the exception of physical activity and sleep. The limited provision of physical activity and sleep data by participants possibly indicates that these aspects, unlike diet, were not viewed as central to the study, despite encouragement to include this information. The minimal study duration was 14 days (cohort B), which may represent a significant time investment for participants. Besides participant’s fatigue, other parameters may have impacted adherence. For example, technical issues such as faulty glucose sensors or omission to scan the sensor and temporary technical issues in the early versions of the food tracking app or the “Food & You” website may at times have contributed to a lower amount of data delivered. However, across both cohorts, data availability was high for most indicators.

Because the “Food & You” study obtained high-resolution dietary data from a mobile app (MyFoodRepo) that was originally created specifically for the study, care needed to be taken to assess the quality of the nutritional data. As other studies have also begun to use the app, the first independent validation study indicated strong data quality^19^. In addition, the fact that every submitted data point on nutrition was reviewed by a study annotator provides additional confidence in the data quality. We observed expected patterns related to weekdays and weekends in terms of timing of food intake, glucose curves, and wake-up and bedtimes at the population level. The high proportion of CGM peaks matching a food intake further suggests that participants logged their food intake appropriately and that the number of missing intakes is expected to be low. In addition, overall intake, and intake of main food groups is in agreement with results reported from a representative sample of the Swiss population.

“Food & You” is the first study that collected food intake via the AI-assisted nutrition tracking app MyFoodRepo, generating an unparalleled dietary high temporal resolution dataset of over 300’000 food dishes with a total of over 45 million kcal. We therefore have a very precise picture of dietary patterns over at least two weeks from over 1000 participants. The data provided by the MyFoodRepo app is primarily based on time-stamped pictures, and thus allows for objective temporal assessment of food intake. The high study completion rates after participants completed the test of the app, combined with the overall positive feedback from post-study surveys indicate that MyFoodRepo was well accepted by participants.

Self-selection bias may have led to a non-representative study population with regard to some particular sociodemographic variables. Based on the exclusion criteria targeting the non pre-diabetic and diabetic population, we expected our study population to lead a healthy lifestyle. Indeed, the high proportion of physically active people who self-report a good health status among the participants points in that direction. Similarly, blood glucose-related indicators such as MAGE and proportion of hypo- and hyperglycemic excursion show no indication of subjects affected by pre-diabetes. Recruitment via social media may have increased the proportion of younger participants with scientific affiinity. Further, the digital nature of the project could have increased the selection of digitally savvy participants. The lack of socio-demographic representation of the “Food & You” study can be overcome through appropriate weighting of the imbalanced strata as conducted in other studies^36^. This adjustment procedure will be crucial for further publications, since it has been shown that socio-demographic factors greatly influence dietary factors^37^. Finally, by excluding pre-diabetic people from the study, we may have filtered out participants with high BMI, since the former is known to be associated with the latter. A Swiss study^38^ on self-reported anthropometric measurements has reported that BMI measurements based on self-reported height and weight are underestimated by a factor of 1.6. The true distribution of BMI observed in the “Food & You” project may therefore be shifted towards higher BMI ranges.

The use of a non-blinded CGM system may be considered a limitation of this study as participants may have adjusted their dietary habits with the intention to control their glucose levels. The decision to use a non-blinded CGM system was motivated by two factors. First, we believed that participants would remain more engaged than with a blinded system. Second, as self-tracking health sensors become more commonly available, future personalized nutrition systems deployed at scale must be designed in the context of full data visibility to participants. Whether such potential self-adjustments in digital cohorts are limited to the short term and initial use is an important question for further research.

Microbiome samples in “Food & You” were sequenced using 16S rRNA, whereas the samples from the other studies to which the microbiome data was compared to used shotgun sequencing. The distinct clustering of “Food & You” and LifeLinesDeep samples, originating from Switzerland and the Netherlands respectively, suggests a potential geographical influence. These geographical differences are likely contributing factors to the observed variations in gut microbiota between the two cohorts. More detailed analyses of the microbiome and its associations with other data collected in the study are ongoing.

Taken together, our results show that collecting a large amount of high-quality data with high study protocol adherence is feasible in the framework of a digital nutrition cohort, opening the path toward large-scale and detailed personalized nutrition studies.

## Data Availability

All data produced in the present study will be made available upon peer-reviewed publication.

## Acknowledgments

We are deeply grateful to the participants of the “Food & You” study. This work was supported by the Kristian Gerhard Jebsen Foundation, the Seerave Foundation, and the Fondation Leenaards. The funders had no role in the design or execution of this study and will have no role in the analyses, interpretation of the data, or decision to submit results. We thank Stéphanie Bonnewyn, Sandra Rüegsegger-Tanner, and Bennan Tong for help with the annotations, as well as Dr Jardena Puder for her clinical guidance. We further thank everyone who helped with various parts of the project, especially Chloé Greub, Leila Munaretto, and Jil Zanolin.

## Author Contributions

Study Design: CA, MS

Technological Development: AB, SC, NF, GH, YJ, DK, PdV

Nutritional Analysis: CA, GR-L, RMS, EU-B

Project Management: CA, SR, TS, HH, MS

Data analysis: HH, VB, DK, RS, LS, MS

Writing: HH, RS, LS, MS

## Data Availability

The data to reproduce the results presented will be made available in a public repository upon publication. We’re happy to share the repository with reviewers during the review process.

## Footnotes

1 https://www.myfoodrepo.org/

2 https://world.openfoodfacts.org/

