## Supplementary Material for "Food & You: A Digital Cohort on Personalized Nutrition"

*Harris Héritier[1], Chloé Allémann[1], Oleksandr Balakiriev[1], Victor Boulanger[1], Sean F. Carroll[1], Noé Froidevaux[1], Germain Hugon[1], Yannis Jaquet[1], Djilani Kebaili[1], Sandra Riccardi[1], Geneviève Rousseau-Leupin[1], Rahel M. Salathé[1], Talia Salzmann[1], Rohan Singh[1], Laura Symul[1,2], Elif Ugurlu-Baud[1], Peter de Verteuil[1], Marcel Salathé[1]*

[1] Digital Epidemiology Lab, School of Life Sciences, School of Computer and Communication Sciences, EPFL, Switzerland

[2] Department of Statistics, Stanford University, USA

##### Supplementary Table 1

Table 1: Inclusion and exclusion criterii for the B and C cohorts of the Food and You study

| Inclusion Criteria | B Cohort | C Cohort |
|---|---|---|
| Aged over 18 | ✓ | ✓ |
| Swiss residency | ✓ | ✓ |
| To own a NFC-enabled smartphone | ✓ | ✓ |
| French German or English speaking | ✓ | ✓ |
| To take part in the project only once | ✓ | ✓ |
| Not in pregnancy | ✓ | ✓ |
| Not in dialysis | ✓ | ✓ |
| Not taking immunosuppressive medication | ✓ | ✓ |
| Not breastfeeding | n.a. | ✓ |
| No antibiotic intake in the last thee months | ✓ | ✓ |
| Absence of chronic gastrointestinal disorder | ✓ | ✓ |
| Absence of active inflammatory or neoplastic disease in the last 3 years | ✓ | ✓ |
| Absence of neuropsychiatric disorder | ✓ | ✓ |
| Absence of myocardial infarction or cerebrovascular accident in the last 6 months | ✓ | ✓ |
| Absence of (pre-) diagnosis for type 1 or 2 Diabetes Mellitus | ✓ | ✓ |
| No hormonal contraceptive intake | n.a. | ✓ |
| No amenorrhea | n.a. | ✓ |
| No (pre-) menopaused | n.a. | ✓ |
| No abortion in the last 3 months | n.a. | ✓ |

### Supplementary Table 2

Table 2: Daily standardized breakfasts in relation to dietary restrictions for both cohorts

| Diet | Days 2-3 (*16-17) | Days 4-5 (*18-19) | Days 6-7 (*20-21) |
|---|---|---|---|
| Normal | 110g white bread | 110g white bread with 30g butter | 50g glucose |
| Non-dairy | 110g white bread | 55g white bread with 55g dark chocolate | 50g glucose |
| Non-gluten | 110g gluten-free white bread | 110g gluten-free white bread with 30g butter | 50g glucose |
| Non-dairy-gluten | 110g gluten-free white bread | 55g gluten-free white bread with 55g dark chocolate | 50g glucose |

* Additional tracking period for cohort C participants

#### Supplementary Table 3

| Category | Energy (kcal) | Meat (g) | Dairy (g) | Water (g) | Fruits and Vegetables > 5 (%) | n Observations |
|---|---|---|---|---|---|---|
| total | 2205.19 (728.13) | 91.99 (111.83) | 124.81 (145.81) | 965.06 (954.96) | 6.81 | 1013 |
| female | 2066.8 (638.03) | 81.63 (101.98) | 116.77 (134.86) | 967.76 (1006.26) | 7.04 | 568 |
| male | 2390.87 (797.05) | 105.9 (122.46) | 135.59 (158.71) | 961.45 (881.56) | 6.52 | 445 |
| 18-34 | 2226.85 (757.97) | 81.21 (113.8) | 121.05 (150.4) | 1017.16 (910.85) | 4.85 | 412 |
| 35-49 | 2228.07 (709.53) | 101.6 (112.6) | 120.34 (138.13) | 948.41 (796.66) | 5.95 | 370 |
| 50-64 | 2131.81 (706.14) | 97.37 (105.85) | 133.74 (148.01) | 913.28 (1294.08) | 9.66 | 207 |
| 65+ | 2076.04 (612.48) | 82.23 (95.8) | 186.8 (148.76) | 754.72 (541.56) | 29.17 | 24 |
| german | 2224.47 (733.05) | 86.18 (109.82) | 132.64 (150.85) | 1079.26 (1087.83) | 8.01 | 549 |
| latin | 2183.18 (721.93) | 98.63 (113.73) | 115.86 (139.32) | 834.69 (755.25) | 5.39 | 464 |
| female.18-34 | 2074.62 (645.9) | 70.64 (99.77) | 108.97 (127.6) | 1003.57 (856.08) | 5.93 | 253 |
| female.35-49 | 2109.16 (626.29) | 91.25 (104.05) | 122.13 (136.06) | 936.19 (732.74) | 5.76 | 191 |
| female.50-64 | 1988.94 (639.18) | 91.48 (101.86) | 121.65 (145.69) | 959.38 (1581.11) | 9.32 | 118 |
| female.65+ | 1801.92 (450.93) | 43.71 (67.55) | 182.75 (155.41) | 624.63 (435.9) | 50.0 | 6 |
| male.18-34 | 2481.73 (857.1) | 98.91 (132.21) | 141.26 (180.56) | 1039.92 (995.77) | 3.14 | 159 |
| male.35-49 | 2362.39 (771.58) | 113.28 (120.51) | 118.32 (140.44) | 962.21 (863.23) | 6.15 | 179 |
| male.50-64 | 2331.43 (746.18) | 105.6 (110.75) | 150.64 (149.67) | 848.86 (718.57) | 10.11 | 89 |
| male.65+ | 2163.9 (632.34) | 94.58 (100.32) | 188.1 (147.05) | 796.42 (566.23) | 22.22 | 18 |

Selected nutritional statistics of the cohort shown as weighted means and standard deviations (in parentheses) of nutrient intake by demographic groupings. Data weighted according to each unique combination of season, day of the week, and individual. Weights were computed as reciprocals of the frequency of each unique combination in the dataset, ensuring a balanced representation of less frequent groupings. Column “Fruits and Vegetables” shows fraction of participants eating at least 5 portions of fruits and vegetables per day, in percent.

### Supplementary Figure 1

**a)**

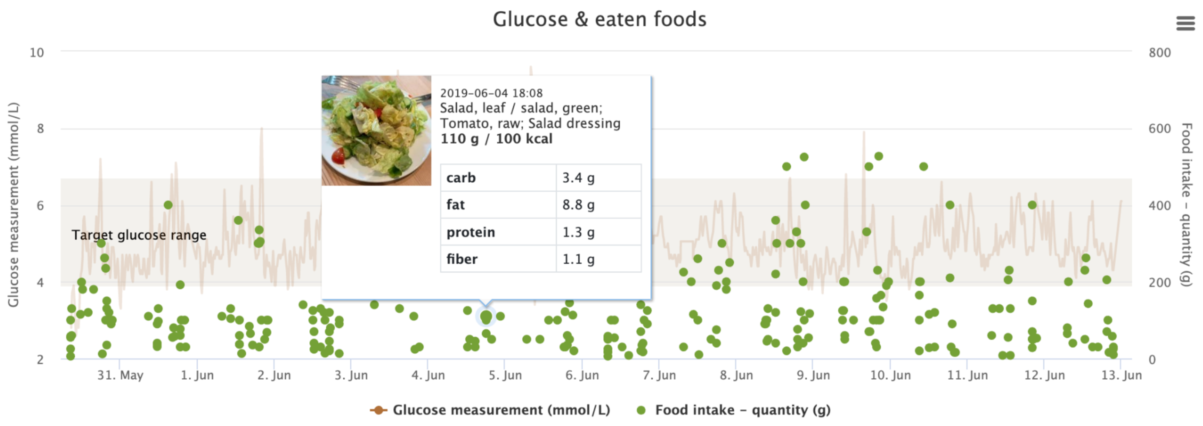

**b)**

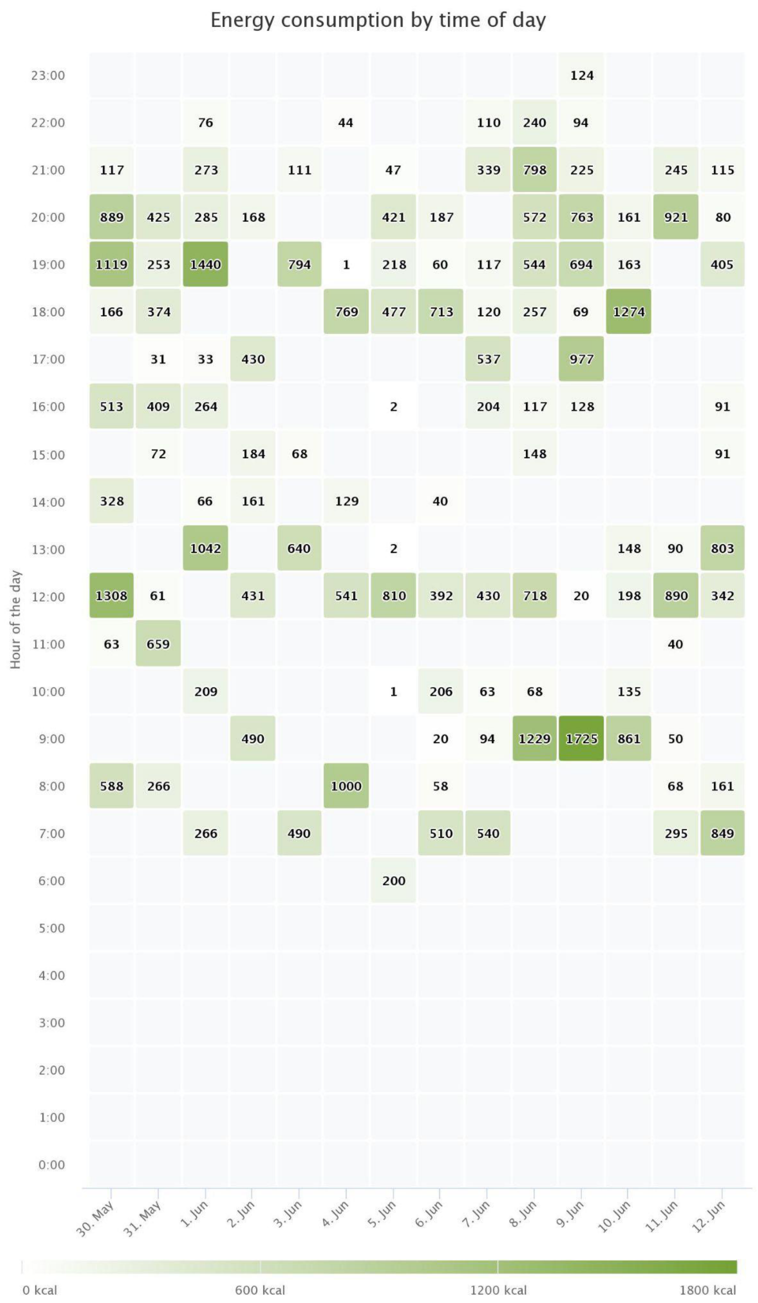


Example of charts shown to participants. a) Glucose response overlayed with annotated dishes. b) Energy consumption by day.

#### Supplementary Figure 2

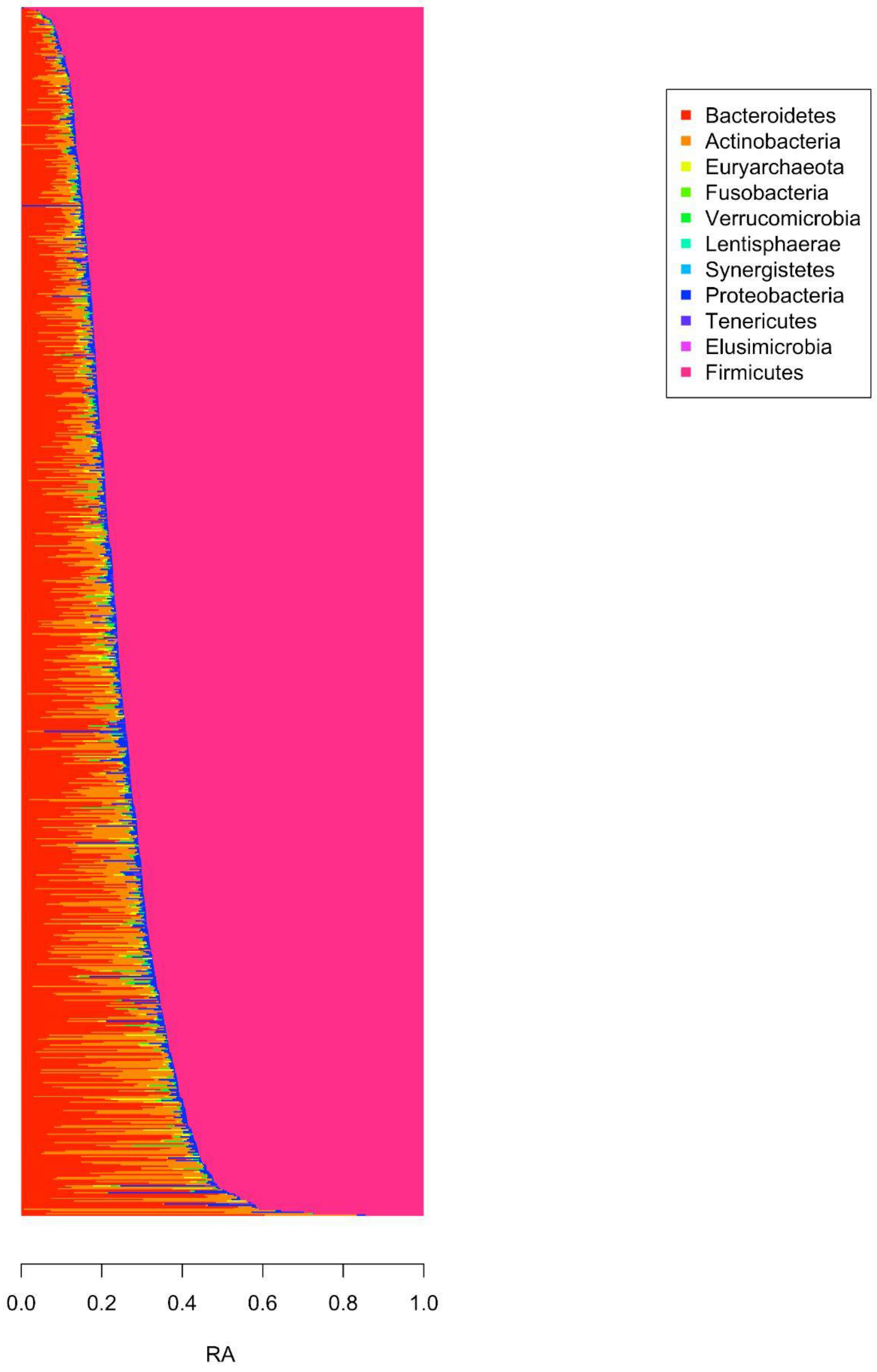


Relative abundance of gut microbiome of “Food & You” participants at phylum level

#### Supplementary Figure 3

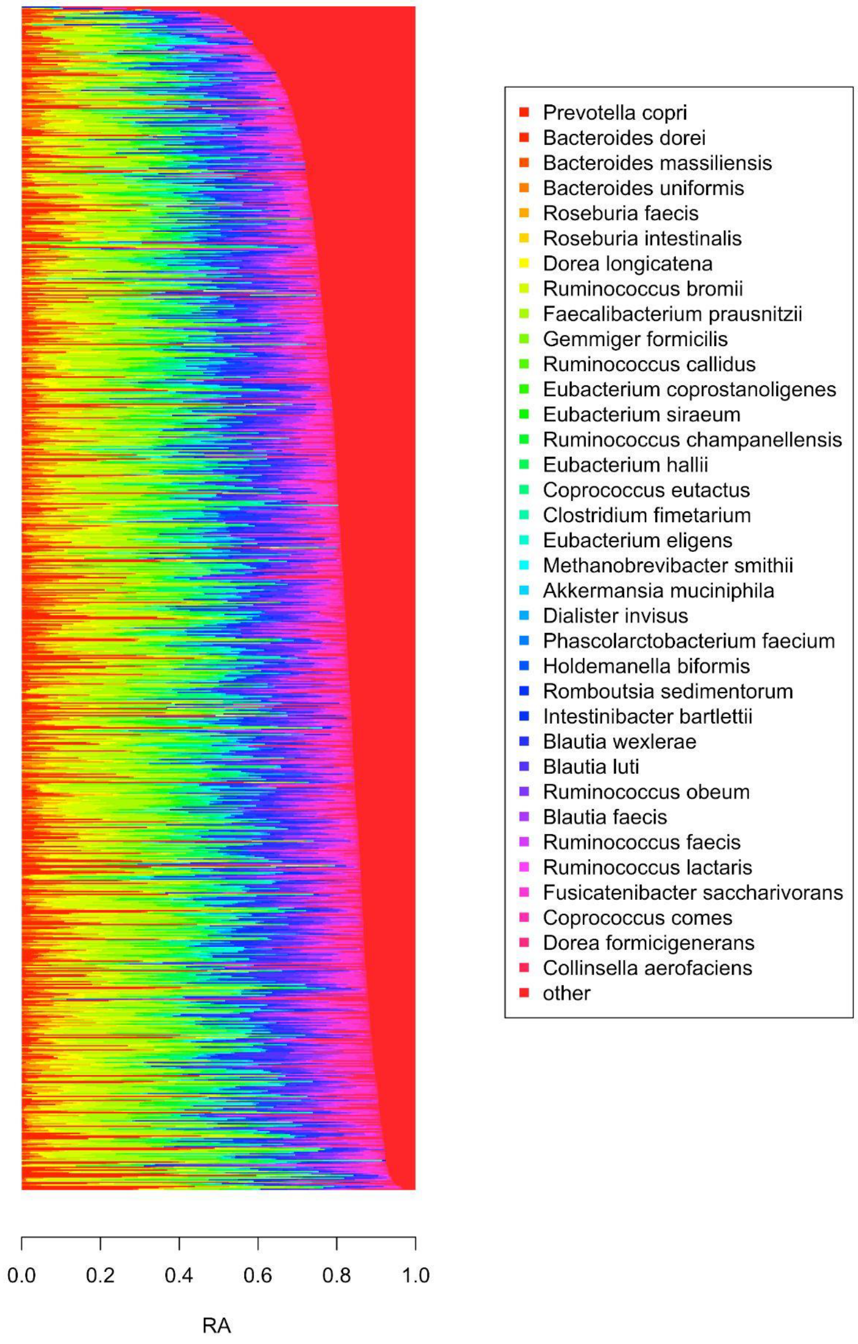


Relative abundance of gut microbiome of “Food & You” participants at species level
